# Evaluate the reliability of the apprenticeship in the first year of medical school: towards a reliable first level ultrasound examination

**DOI:** 10.1101/2022.04.08.21259596

**Authors:** Michela Cristina Turci, Massimo Tassoni, Florida Gajtani, Antonino Michele Previtera, Alberto Magenta-Biasina, Elia Mario Biganzoli, Graziano Serrao

## Abstract

**Objectives:** Our aim was to motivate apprentices’ sonographer needs, to appraise their own measurements, to reduce inconsistencies within and between operators. Deep knowledge of ultrasound sectional anatomy is mandatory for an appropriate performance.

**Methods:** In three different weekdays, 3 sonographer apprentices (**rater**), randomly selected from a cohort of San Paolo Medical School first year students participated in vertically integrated study of living anatomy through ultrasound examination, repeated lumbar multifidus cross-sections scans on 6 subjects at lumbar level. The Agreement R package 0.8-1 was used to monitored the performances of each apprentice.

**Results:** The agreement (CCC_intra_ 0.6749; CCC_inter_ 0.556; CCC_total_ is 0.5438) was further from least acceptable CCC of 0.92-0.95. The precision indices (precision_intra_ 0.6749; _inter_ 0.801; _total_0.6274) were unsatisfactory, while the accuracy was high (0.9889 to 0.9913). The same occurred for the agreement on rater performances comparisons, where readings were high accurate (0.9537 to 0.9733) but moderately precise (0.7927 to 0.8895), not interchangeable TIR (1.173) but without rater supremacy. IIR (_r1 vs r2_ 1.104, _r1 vs r3_ 1.015, _r2 vs r3_ 0.92) 95% confidence limits.

**Conclusions:** Apprentices were not reliable, repeatable, interchangeable. The weak link in the method seemed to be cultural weakness on vivo imaging morphologies, qualitative and quantitative measurement procedure on elementary statistical processing.

## Introduction

Moved by evidence, teachers must work hardly to transform excellence into routine performances. Handle echography represents a key toll that allows an immediate correlation between imaging findings and clinical data, which will improve the management of prime care supply at the patient’s home. The fast advancement on telemedicine, the increased use and the decreasing instrument costs lead to a great demand for appropriate education toward a sturdy echo’s competence [1-5]. Echo continuously improves the instrumental semiotics potentials by making more anatomic structures visible. Since 2007 at San Paolo Medical School, an ultrasound learning resource was located within the anatomy area [6]. The focus of this educational training would be the capacity to forge a physician able to execute a first level ultrasound examination (**FLUx**). Often in clinical practice, only one measurement per patient is performed, hence before getting down to the data analysis you want to ensure that they are not contaminated by assessed procedure/factors.

Our aim was to verify apprentice’s judgement validity and tune learning path.

## Methods

Ultrasonic method is commonly described as largely employable, pliant, and non-invasive [6,7]. In 2009, San Paolo Medical School Didactic College planed vertically integrated study of the living body anatomy through peer physical and ultrasound examination. Core group of faculty experts in ultrasound developed a standardized curriculum that was presented to the student body. Faculty-supported facility for medical students’ independent education projects allowed each student to be alternately examine and examiner [8]. To evaluate the competences acquired and tune the training on academic year 2019-2020, 3 women and 3 men (ranging in age 19 to 22±1.27yr, and in BMI 19.0 to 25.1±2.42 kg×m^-2^) healthy apprentices, ruffled among participants in the ultrasound living anatomy course took part in the present study as examines, but only 3 as examiners too. The Ethics Committee of the University of Milan examined and issued, an ex post favourable opinion on the project (03/28/2022 advisory 31/22).

### Scanning procedure

An experienced physiatrist (**AMP**) evaluated each drawn student to exclude any sequels that could alter the region of interest. A multi-frequency probe equipped ultrasound machine B-mode Real Time (Logic QE, Medical Systems, Milwaukee, WI, USA) was used. A convex probe at 5 MHz emission was employed [9,10], 3 apprentices (**raters**), under conditions as close as possible to the clinical daily routine, performed ultrasound scans then run measurement/rating process.

Following the procedure commonly recommended in literature, each rater proceeded by steps. To flatting down the lumbar lordosis the subject lied prone with a pillow under the pubic bones and feet off the bed, arms along her/his hips and head turned toward the most comfortable side (**Fig. 1**). To identify the spinous processes of L4-L5 the rater aligned each of her/his index fingers with the correspondent iliac crest and extended the thumbs towards the spinous processes of the vertebral column (**Fig.2**) [11]. To confirm the level found by palpation, each rater verifies by ultrasound scan [12]. In order to allow the vertebral lamina, the multifidus muscle was scanned longitudinally (**Fig. 3**).

**Figure 1.**
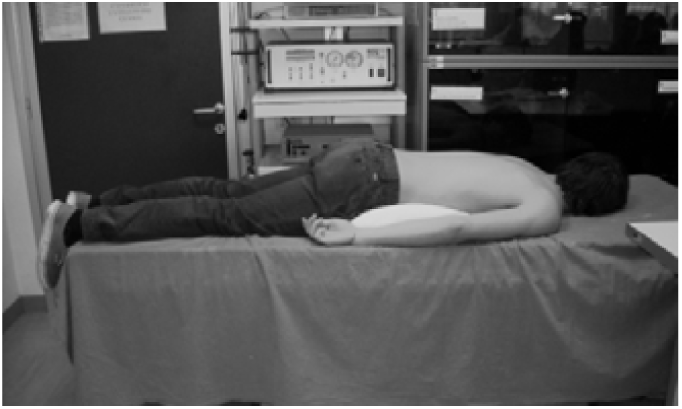
Subject position.

**Figure 2.**
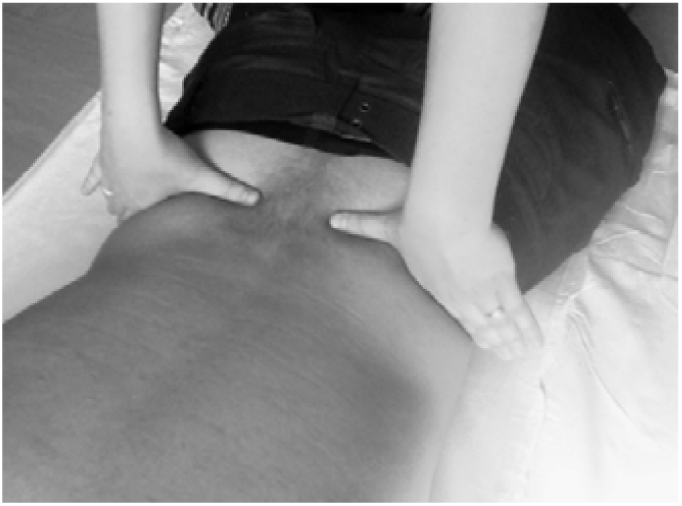
Identification of the spinous processes.

**Figure 3.**
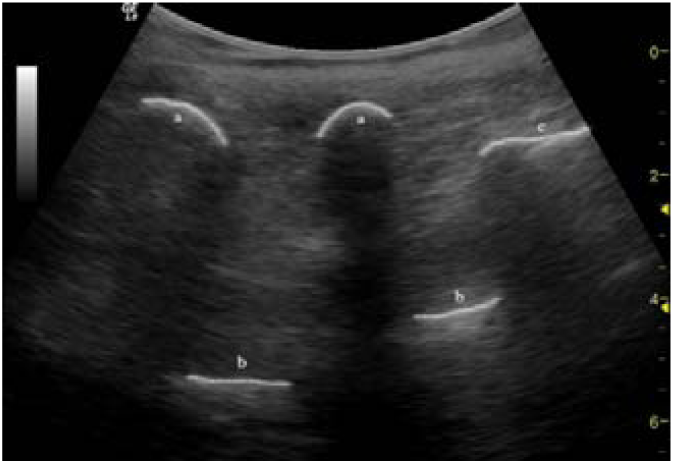
Lumbar longitudinal scan btween L4 and sacrum a: apex of the spinous process, b: yellow ligament, c: median sacral crest.

Thereafter in order to be transverse correctly aligned, the rater performed a 90° rotation of the probe. The discrimination of lateral edges of multifidus out of the surrounding muscles was the most difficult part of the scanning protocol. Each subject was requested of slightly lifting the ipsilateral lower limb from the couch; the identification of that lateral edge was so favored. After the contraction, before measuring the muscle diameters, the rater waited a few seconds to settle the rest position [13,14]. After having identified the multifidus, the rater moved the probe to the left and then to the right while tracing antero-posterior (r/lAP) and latero-lateral diameter (r/lLL) assessments. On frozen images, the AP diameter, from the lamina to the inner edge of the upper edge of the multifidus, and the LL diameter, corresponding to the maximum width from the lateral edge of the multifidus to the spine, were marked out. Each rater established independently the positioning of the subject and the anatomical landmarks. Raters were not aware to other rater’s judgments. In accord with Watson statements [15,16], 3 repeated measurements are usually considered appropriate, hence on different weekdays, the procedure was 3 times repeated, on 6 subjects, at five vertebral levels, on each side and therefore 180 scans generated 360 diameters.

### Statistical analysis

Reliable and accurate measurements have a common theme: assessing closeness (agreement) between observations. Because there are 3 raters involved, one may like assessing the intra, inter and the total agreement with replicated readings produced by different observers. A meaningful statistic to measure the agreement of observations has been the mean squared deviation (**MSD**). The *Concordance Correlation Coefficient* (**CCC**), a scaled index proposed by Lin [17,18], translates the MSD into a combine’s coefficients of precision (degree of refinement) and accuracy (degree of total displacement) that can be used to compare the differences between operators and sessions. To esteem multiple observers with replications, where none of the observers is treated as reference, Barnhart [19] proposed CCCr_otal_, CCC_inter_, and CCC_intra_ series of indices. Another intuitive agreement index is a measure that captures a large proportion of data within a settled maximum acceptable absolute difference between two observers’ readings. This probability is called *coverage probability* (**CP**). On the other hand, if we set a predetermined coverage probability, we can find the boundary so that the probability of absolute difference is less than the boundary itself. This boundary called *total deviation index* (**TDI**) is proportional to the square root of the MSD and it is a performance measurement. A satisfactory agreement may require a large CP or equivalently a small TDI [18,20,21].

When multiple raters are available with replicates, we are often interested if raters are interchangeable. The *Total-Intra Ratio* (**TIR**) assesses comparative agreement indices for the interchangeability between multiple raters with multiple readings. The scale criterion means that the MSD_total-raters_ cannot be more than a predefined value of the MSD_intra-rater_, or TIR_R_ < predefined value, with 95% confidence. In addition, we compared the intra rater precision by computing MSD_intra1_2_/MSD_intra3_, MSD_intra1_3_/MSD_intra2_, and MSD_intra2_3_/MSD_intra1_ or *Intra-Intra Ratio* (**IIR**). A 100(1 − α/2) % confidence interval for IIR is computed based on the log-transformed IIR estimates and claim the superiority or inferiority if the upper or lower limit is less than or greater than 1.0 [22]. All index was computed by Agreement R package 0.8 [23].

## Results

**Table 1** shows the multifidus cross section AP and LL diameters measurements (mm) relieved by each rater, for each subject and lumbar level.

**Table 1.** AP e LL diameters values (mm) of the multifidus relieved by each rater are shown for each subject, lumbar level and repeat (t1,2,3).

| subject | level | right anteropost |  |  |  |  |  |  |  |  | left anteropost |  |  |  |  |  |  |  |  | right laterolateral |  |  |  |  |  |  |  |  | left laterolateral |  |  |  |  |  |  |  |  |
| --- | --- | --- | --- | --- | --- | --- | --- | --- | --- | --- | --- | --- | --- | --- | --- | --- | --- | --- | --- | --- | --- | --- | --- | --- | --- | --- | --- | --- | --- | --- | --- | --- | --- | --- | --- | --- | --- |
|  |  | rater1t1 | rater1t2 | rater1t3 | rater2t1 | rater2t2 | rater2t3 | rater3t1 | rater3t2 | rater3t3 | rater1t1 | rater1t2 | rater1t3 | rater2t1 | rater2t2 | rater2t3 | rater3t1 | rater3t2 | rater3t3 | rater1t1 | rater1t2 | rater1t3 | rater2t1 | rater2t2 | rater2t3 | rater3t1 | rater3t2 | rater3t3 | rater1t1 | rater1t2 | rater1t3 | rater2t1 | rater2t2 | rater2t3 | rater3t1 | rater3t2 | rater3t3 |
| 1 | L1 | 154 | 158 | 177 | 125 | 159 | 160 | 179 | 146 | 141 | 140 | 149 | 163 | 110 | 151 | 149 | 153 | 139 | 137 | 206 | 216 | 192 | 179 | 168 | 199 | 318 | 163 | 98 | 182 | 182 | 163 | 193 | 220 | 230 | 291 | 208 | 163 |
| 2 | L1 | 231 | 198 | 218 | 161 | 219 | 197 | 193 | 176 | 155 | 190 | 218 | 215 | 159 | 175 | 202 | 168 | 166 | 159 | 216 | 210 | 156 | 186 | 163 | 172 | 314 | 182 | 188 | 174 | 181 | 148 | 135 | 202 | 152 | 296 | 202 | 210 |
| 3 | L1 | 213 | 219 | 252 | 190 | 165 | 194 | 159 | 162 | 146 | 193 | 227 | 196 | 191 | 168 | 189 | 159 | 152 | 157 | 326 | 227 | 204 | 192 | 188 | 233 | 296 | 202 | 126 | 286 | 223 | 181 | 177 | 191 | 220 | 186 | 185 | 168 |
| 4 | L1 | 215 | 168 | 191 | 163 | 199 | 143 | 142 | 162 | 160 | 175 | 132 | 188 | 159 | 188 | 151 | 148 | 141 | 157 | 177 | 244 | 204 | 161 | 226 | 202 | 95 | 124 | 106 | 140 | 226 | 182 | 172 | 189 | 150 | 95 | 118 | 103 |
| 5 | L1 | 180 | 129 | 169 | 164 | 165 | 141 | 149 | 123 | 121 | 177 | 142 | 167 | 115 | 121 | 158 | 132 | 129 | 104 | 217 | 208 | 154 | 136 | 243 | 187 | 156 | 171 | 159 | 203 | 213 | 171 | 139 | 219 | 197 | 187 | 169 | 152 |
| 6 | L1 | 192 | 173 | 202 | 169 | 164 | 202 | 198 | 137 | 180 | 236 | 172 | 195 | 168 | 156 | 212 | 173 | 152 | 188 | 148 | 247 | 160 | 207 | 257 | 221 | 159 | 152 | 163 | 168 | 209 | 277 | 179 | 218 | 174 | 229 | 229 | 205 |
| 1 | L2 | 210 | 226 | 165 | 199 | 222 | 192 | 226 | 202 | 173 | 237 | 248 | 216 | 208 | 239 | 184 | 218 | 183 | 173 | 377 | 254 | 223 | 350 | 261 | 235 | 354 | 194 | 143 | 345 | 279 | 230 | 421 | 251 | 224 | 408 | 263 | 143 |
| 2 | L2 | 154 | 201 | 203 | 127 | 223 | 215 | 202 | 205 | 188 | 156 | 233 | 257 | 118 | 262 | 213 | 205 | 166 | 168 | 177 | 327 | 188 | 174 | 251 | 195 | 285 | 197 | 241 | 174 | 318 | 158 | 179 | 353 | 202 | 283 | 260 | 235 |
| 3 | L2 | 219 | 216 | 206 | 155 | 192 | 213 | 238 | 171 | 207 | 191 | 156 | 247 | 153 | 179 | 270 | 213 | 143 | 176 | 215 | 187 | 236 | 159 | 253 | 182 | 269 | 123 | 152 | 219 | 176 | 191 | 179 | 173 | 190 | 261 | 126 | 123 |
| 4 | L2 | 262 | 182 | 177 | 210 | 182 | 187 | 257 | 140 | 188 | 250 | 175 | 212 | 247 | 164 | 187 | 247 | 165 | 160 | 386 | 215 | 276 | 332 | 255 | 184 | 309 | 155 | 132 | 321 | 179 | 211 | 278 | 249 | 182 | 405 | 143 | 127 |
| 5 | L2 | 183 | 224 | 156 | 193 | 185 | 172 | 163 | 187 | 129 | 211 | 196 | 203 | 161 | 210 | 195 | 132 | 173 | 135 | 202 | 195 | 271 | 161 | 147 | 218 | 89 | 159 | 142 | 206 | 168 | 168 | 202 | 173 | 164 | 101 | 159 | 145 |
| 6 | L2 | 223 | 208 | 216 | 189 | 201 | 202 | 160 | 189 | 199 | 183 | 261 | 185 | 163 | 176 | 210 | 176 | 194 | 171 | 202 | 213 | 245 | 182 | 263 | 237 | 176 | 192 | 225 | 188 | 220 | 277 | 166 | 248 | 207 | 173 | 242 | 235 |
| 1 | L3 | 221 | 237 | 171 | 177 | 217 | 238 | 198 | 256 | 185 | 217 | 237 | 181 | 228 | 263 | 255 | 159 | 222 | 168 | 247 | 346 | 230 | 213 | 268 | 218 | 215 | 241 | 114 | 194 | 310 | 243 | 167 | 304 | 249 | 199 | 235 | 182 |
| 2 | L3 | 163 | 229 | 260 | 134 | 257 | 271 | 181 | 188 | 222 | 187 | 248 | 282 | 158 | 233 | 268 | 165 | 182 | 224 | 222 | 335 | 254 | 179 | 350 | 289 | 283 | 168 | 258 | 190 | 401 | 226 | 253 | 256 | 263 | 306 | 303 | 242 |
| 3 | L3 | 224 | 187 | 258 | 185 | 191 | 282 | 263 | 204 | 187 | 247 | 204 | 218 | 219 | 209 | 276 | 265 | 193 | 202 | 302 | 304 | 254 | 245 | 213 | 294 | 412 | 131 | 131 | 273 | 266 | 227 | 161 | 203 | 238 | 385 | 212 | 233 |
| 4 | L3 | 267 | 133 | 199 | 252 | 168 | 207 | 245 | 163 | 204 | 260 | 171 | 196 | 248 | 193 | 201 | 260 | 182 | 200 | 376 | 263 | 323 | 317 | 251 | 214 | 335 | 219 | 187 | 355 | 228 | 254 | 206 | 280 | 258 | 378 | 219 | 193 |
| 5 | L3 | 175 | 199 | 152 | 207 | 230 | 165 | 188 | 180 | 141 | 225 | 195 | 163 | 214 | 212 | 190 | 185 | 176 | 168 | 221 | 251 | 288 | 249 | 177 | 212 | 154 | 171 | 140 | 221 | 283 | 289 | 352 | 188 | 179 | 190 | 183 | 143 |
| 6 | L3 | 268 | 187 | 263 | 192 | 193 | 230 | 182 | 177 | 205 | 208 | 228 | 224 | 195 | 223 | 223 | 202 | 251 | 213 | 241 | 286 | 299 | 223 | 305 | 240 | 227 | 294 | 228 | 314 | 289 | 248 | 234 | 327 | 287 | 227 | 346 | 253 |
| 1 | L4 | 257 | 181 | 204 | 270 | 193 | 230 | 179 | 246 | 229 | 236 | 210 | 205 | 203 | 159 | 223 | 218 | 328 | 233 | 313 | 372 | 296 | 230 | 288 | 283 | 272 | 330 | 278 | 245 | 388 | 290 | 267 | 256 | 321 | 246 | 306 | 286 |
| 2 | L4 | 185 | 261 | 275 | 139 | 248 | 260 | 223 | 291 | 241 | 187 | 240 | 235 | 136 | 230 | 257 | 199 | 326 | 258 | 253 | 341 | 292 | 193 | 336 | 329 | 291 | 409 | 333 | 215 | 244 | 252 | 200 | 208 | 349 | 370 | 359 | 338 |
| 3 | L4 | 259 | 212 | 257 | 206 | 232 | 293 | 257 | 270 | 297 | 243 | 233 | 248 | 163 | 236 | 261 | 321 | 260 | 308 | 335 | 379 | 294 | 321 | 382 | 394 | 377 | 300 | 317 | 229 | 354 | 294 | 177 | 326 | 288 | 341 | 330 | 353 |
| 4 | L4 | 250 | 177 | 216 | 255 | 171 | 240 | 262 | 205 | 247 | 265 | 217 | 232 | 266 | 201 | 254 | 266 | 205 | 255 | 387 | 248 | 363 | 432 | 229 | 311 | 344 | 309 | 333 | 359 | 267 | 307 | 343 | 257 | 329 | 417 | 341 | 314 |
| 5 | L4 | 253 | 257 | 181 | 186 | 238 | 161 | 257 | 213 | 177 | 242 | 215 | 213 | 201 | 243 | 131 | 254 | 191 | 187 | 176 | 332 | 333 | 229 | 227 | 199 | 316 | 211 | 168 | 173 | 411 | 326 | 237 | 243 | 200 | 302 | 235 | 182 |
| 6 | L4 | 261 | 200 | 232 | 161 | 235 | 227 | 218 | 228 | 286 | 240 | 198 | 233 | 157 | 205 | 255 | 193 | 269 | 297 | 247 | 308 | 275 | 233 | 305 | 321 | 364 | 300 | 395 | 203 | 287 | 286 | 262 | 344 | 349 | 293 | 351 | 320 |
| 1 | L5 | 238 | 202 | 224 | 259 | 217 | 227 | 261 | 258 | 218 | 271 | 179 | 187 | 267 | 203 | 221 | 286 | 255 | 211 | 374 | 397 | 319 | 351 | 337 | 336 | 311 | 426 | 331 | 370 | 396 | 282 | 341 | 286 | 351 | 307 | 443 | 283 |
| 2 | L5 | 190 | 263 | 229 | 149 | 248 | 229 | 193 | 256 | 256 | 199 | 260 | 247 | 165 | 236 | 238 | 185 | 266 | 260 | 272 | 362 | 369 | 204 | 382 | 374 | 403 | 320 | 392 | 277 | 327 | 325 | 219 | 360 | 376 | 299 | 343 | 376 |
| 3 | L5 | 219 | 191 | 237 | 234 | 161 | 255 | 230 | 247 | 264 | 252 | 207 | 252 | 198 | 215 | 271 | 241 | 256 | 269 | 352 | 363 | 344 | 382 | 216 | 366 | 445 | 347 | 343 | 327 | 370 | 322 | 252 | 279 | 329 | 445 | 334 | 339 |
| 4 | L5 | 262 | 138 | 212 | 237 | 170 | 216 | 261 | 191 | 242 | 239 | 137 | 224 | 249 | 174 | 202 | 246 | 179 | 253 | 391 | 271 | 359 | 353 | 347 | 193 | 332 | 364 | 353 | 354 | 291 | 367 | 354 | 280 | 201 | 336 | 423 | 359 |
| 5 | L5 | 194 | 201 | 194 | 221 | 290 | 115 | 223 | 280 | 199 | 210 | 251 | 196 | 261 | 285 | 125 | 241 | 275 | 190 | 202 | 359 | 371 | 257 | 296 | 188 | 344 | 326 | 384 | 221 | 316 | 316 | 229 | 320 | 180 | 344 | 330 | 350 |
| 6 | L5 | 256 | 201 | 165 | 249 | 205 | 207 | 201 | 230 | 241 | 229 | 238 | 204 | 219 | 238 | 235 | 177 | 225 | 236 | 268 | 376 | 398 | 326 | 374 | 361 | 342 | 347 | 345 | 273 | 413 | 407 | 306 | 416 | 417 | 391 | 387 | 382 |

Based on multifidus assessments conducted by experts in the field found out on literature in terms of agreement, we set a within-rater deviation not more than 30% of the total deviation, and inter-rater deviation not more than 50% of the total deviation. These translated into a least acceptable CCC_intra_ of 0.91 (1 – 0,3^2^), and a least acceptable CCC_inter_ of 0.75 (1 – 0,5^2^).

The CCC estimates are shown on **Table 2**.

**Table 2.** CCC estimates (see text for details).

| Statistics | Estimate | 95% CI |
| --- | --- | --- |
| CCC<sub>intra</sub> | 0.674 | 0.597 |
| CCC<sub>inter</sub> | 0.792 | 0.732 |
| CCC<sub>total</sub> | 0.621 | 0.627 |
| Precision<sub>intra</sub> | 0.674 | 0.597 |
| Precision<sub>inter</sub> | 0.801 | 0.744 |
| Precision<sub>total</sub> | 0.627 | 0.550 |
| Accuracy<sub>intra</sub> |  |  |
| Accuracy<sub>inter</sub> | 0.988 | 0.980 |
| Accuracy<sub>total</sub> | 0.991 | 0.984 |

**Table 3.** Rater performances (see text for details).

| Statistics | Estimate | 95% CI | Allowance | RBS |
| --- | --- | --- | --- | --- |
| TDI<sub>.95 r1-2</sub> | 5.573 | 6.19 | 5.4 |  |
| TDI<sub>.95 r1-3</sub> | 8.494 | 9.453 | 5.4 |  |
| TDI<sub>.95 r2-3</sub> | 8.608 | 9.581 | 5.4 |  |
| CP<sub>54 r1-2</sub> | 0.941 | 0.908 | 0.95 | 0.234 |
| CP<sub>54 r1-3</sub> | 0.783 | 0.730 | 0.95 | 0.03 |
| CP<sub>54 r2-3</sub> | 0.777 | 0.723 | 0.95 | 0.01 |
| TIR<sub>r1-2-3 vs all</sub> | 1.173 | 1.72 | 1.5 |  |
| IIR<sub>r1 vs r2</sub> | 1.104 | (0.842,1.447) | 1 |  |
| IIR<sub>r1 vs r3</sub> | 1.015 | (0.687,1.499) |  |  |
| IIR<sub>r2 vs r3</sub> | 0.92 | (0.633,1.336) |  |  |

RBS relative bias squared.

The CCC_intra_ was estimated to be 0.6749, which means a within-sample deviation is about 57 % √ (1 – 0.6749) of the total deviations. The *CCC*_*inter*_ was estimated to be 0.7921, which means a within-sample deviation is about √ (1 – 0.791) 63% of the total deviations [17]. The *CCC*_*tota*l_ was estimated to be 0.6219, which means for individual observations from different rater, the within-sample deviation is about 61% of the total deviations. These findings are consistent with moderate precision (0.6749_intra_, 0.8_inter,_ 0.6274_total_), but high accuracy (0.9889_inte_ to 0.9913 _total_) ever larger than 0.95 and 0.91, respectively. Performance analysis of raters is shown in **Table 4**.

The agreement was moderate and further from least acceptable CCC of 0.91-0.95. This finding agrees with the data presented in **Fig. 4a-c**, where readings were evenly scattered around the 45° line (high accuracy 0.9537 to 0.9733) but not tightly scattered (moderate precision 0.7927 to 0.8895). The plots indicated that score differences tended to be more dispersed for large value correspondent to a lower lumbar level.

**Fig 4.**
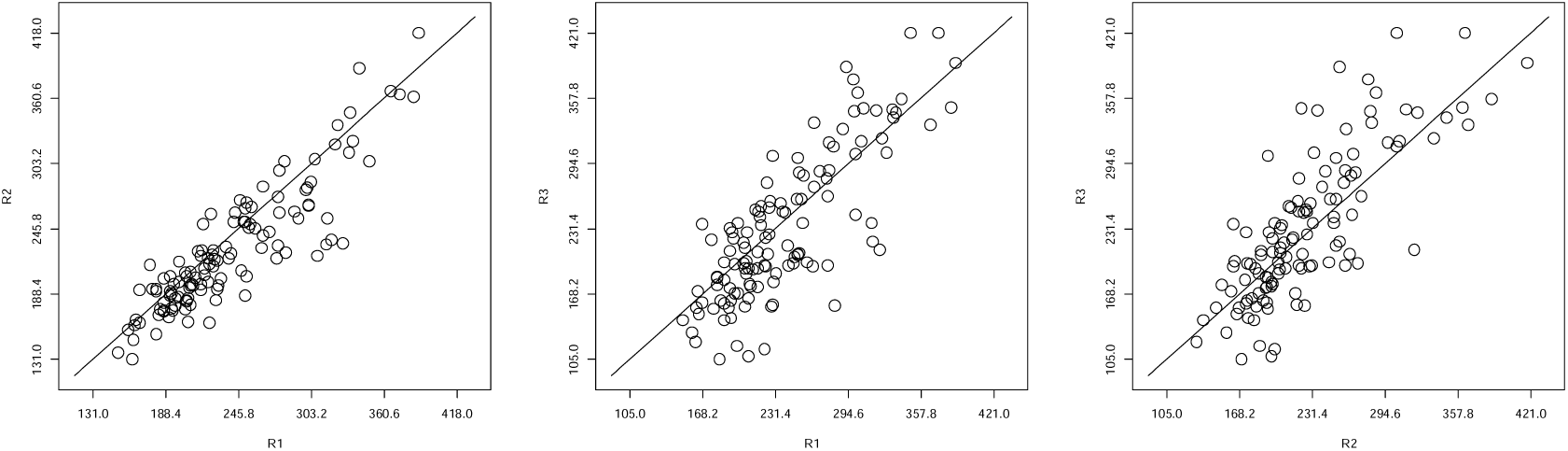
Inter-rater (R) agreement: a R1vsR2; b R1vsR3; c R2vsR3.

In terms of TDI and CP indices, the least acceptable agreement was set as having at least 95% pair’s observations within 5.4 mm (allowance, equal to 20% of all measurements mean), independently to rater. The TDI_0.95r1r2_ estimate was 5.573 mm, close to 5.4 mm target values, which means based on the average readings, 95% of the readings are within 5.573 mm of their replicate readings from the other rater. The TDI_0.95r1r3_ estimate was 8.494 mm, and the TDI_0.95r2r3_ estimate was 8.608 mm. The one-sided upper confidence limit for TDI_0.95r1r2_ was 6.19mm, TDI_0.95r1r3_ 9.453mm, and TDI_0.95r2r3_ 9.581mm all larger than target values. The CP_54r1r2_ estimate was 0.9418, which means that 94% of observations were within their target values from the other rater. The one-sided lower confidence limit for CP_54r1r2_ was 0.9170, smaller than 0.95. The CP_54r1r3_ estimate was 0.7835 and CP_54r2r3_ 0.7773 which means that 78% of observations are within 5.4 mm. The one-sided lower confidence limit for CP_54r1r3_ was 0.7301, and for CP_54r2r3_ was 0.7236 both smaller than 0.95.

TIR _r1-2-3 vs all_ the ratio between average of the total MSD of all raters relative to the average of intra MSD of all raters, was estimated to be 1.173, with a 95% upper confidence limit of 1.72. If we consider that a precision deviation of more than 50% (1.5 allowance) is clinically not acceptable, we cannot claim raters’ interchangeability. The IIR was estimated to be IIR_r1vsr2_ 1.04, IIR _r1vsr3_ 1.015 and IIR _r2vsr3_ 0.92. If we refer at the individual bioequivalence FDA criterion, that the ratio of geometric means between readings differences must lie between 0.8 and 1.25, IIR upper/lower 95% interval {_r1 vs r2_ (0.842, 1.447), _r1 vs r3_ (0.687, 1.499), _r2 vs r3_ (0.633, 1.336)} indicated no rater precision supremacy.

## Discussion

The study confirmed the ultrasound procedure as an operator dependent technique that is prone to variable measurements. Each considered parameter (rater and replicates) induced bias. Each rater proved to be accurate but not sufficiently precise.

Lacking intra-operator and inter-operator reliability made impossible to compare data obtained by different raters of the same department or in different groups and centers, and also frustrated a complete tuning of correct guidelines. Because errors are inherent in every measurement procedure, one must ensure the magnitude of the measurement agreement [18,24,25]. In clinical practice measurements are detect to take a decision, consequently magnitude of acceptable differences between ratings, it depends on clinical decision consequences. No universal standard for an acceptable confidence interval is currently available, but most experts consider range values 0.8-1.25 as supporting good reliability. A confidence interval for the difference between measurement results indicates the smallest detectable difference (**SDD**) [26-29]. With repeated rating, any change outside these boundaries can be considered a true change in the entity being assessed. The smallest detectable difference it is a useful starting point in determining what is a minimum clinically important difference (MCID) in values handle.

When measurements show evidence of lack of agreement, we need to address the sources of the deficiencies. According to Alvan Feinstein [30] for gaining insight into the “*clinimetric”* property, we should reinforce apprentice’s attention on measurements methodologic discipline quality of the measurement [31,32]. To enable assessment in routine care, we’ll beckon worth on agreement, reliability, repeatability, and reproducibility. Because of that, it is essential to reinforce statistical skill in medical training too.

## Conclusions

The cohort sampled was limited but indicative to same training adjustments. Future efforts could focus on tear down “observational errors”. In order to improve rater’s performance, and promote inter-rater reliability, it should be considerers a number of strategies: training on imaging morphologies, nurture feedback to those who have shown a low reproducibility, reinforce protocols. All these methods used as a screening quality, self-evaluation check, and calibration experience encourage raters to achieve 0.8-1.25 range values and gaining smallest detectable difference validity [27,33].

## Data Availability

All data generated or analysed during this study are included in this published article.

